# Length of hospital stay and associated factors among adult surgical patients admitted to a surgical ward in Amhara Regional State Comprehensive Specialized Hospitals, Ethiopia

**DOI:** 10.1101/2023.12.07.23299680

**Authors:** Habtamu Hurisa Dadi, Netsanet Habte, Yenework Mulu, Yabibal Asfaw

## Abstract

**Introduction:** Hospitals across the country are experiencing a rise in the length of hospital stays, ranging from 2% to 14%. As a result, patients who remain hospitalized for a prolonged period are three times more likely to suffer in-hospital deaths. Therefore, identifying contributing factors for prolonged hospital stays enhances the ability to improve services and the quality of patient care. However, there is limited documented evidence in Ethiopia as well as in the study area about factors associated with prolonged hospital stays among surgical inpatients.

**Objective:** This study aimed to assess the length of hospital stay and associated factors among adult surgical patients admitted to a surgical ward in Amhara Regional State Comprehensive Specialized Hospitals, Ethiopia, 2023.

**Methods:** An institutional-based cross-sectional study was conducted among 452 adult surgical patients from April 17 to May 22, 2023. Data were collected based on a pretested, structured interviewer-administered questionnaire, patient chart review, and direct measurement. Study participants were selected using a systematic random sampling technique. The collected data were cleaned, entered into EpiData 4.6.0 and exported to STATA version 14 for analysis. Binary logistic regression analysis was used. Variables with a p value < 0.05 in the multivariable logistic regression analysis were considered statistically significant.

**Results:** In the current study, the prevalence of prolonged hospital stay was 26.5% (95% CI: 22.7–30.8). Patients referred from another public health institution (AOR = 2.46; 95% CI: 1.09, 5.57), hospital-acquired pneumonia (AOR = 3.18; 95% CI: 1.28, 7.89), duration of surgery ≥110 minutes (AOR = 2.48; 95% CI: 1.25, 4.91), and preoperative anemia (AOR = 3.37; 95% CI: 1.88, 6.04) were factors associated with prolonged hospital stays.

**Conclusion:** This study found a significant proportion of prolonged hospital stays. Source of referral, preoperative anemia, duration of surgery, and hospital-acquired pneumonia were factors associated with a prolonged hospital stay. Strengthening the established information system among hospitals when referring patients and early screening and treating anemia upon admission to hospitals can reduce the length of stays.

## Introduction

Hospital admission is essential for improving patient care and providing them with inpatient services, which can help save lives [1]. In fact, hospitals play a vital role in the delivery of healthcare services, and as a result, they significantly affect the efficiency of a health system [2]. Length of stay (LOS) is one of the markers of hospital indicators that is currently utilized regularly to evaluate hospital efficiency and is defined as the date between admission and discharge [3–5]. However, prolonged LOS is defined as a hospitalized stay that is longer than expected for particular treatments [6,7].

Every year, more than 143 million people in low- and middle-income countries undergo surgical procedures, which places a substantial strain on the health care system [8]. As a result, hospitals across the country are experiencing an increase in the length of hospital stays. Just 2% of patients in 2012 had hospital stays longer than 21 days; now, they account for 14% of hospital days and cost more than $20 billion a year [9]. In 2022, patients’ median hospitalizations would be nearly 19% higher than they were in 2019, according to statistics from the healthcare consulting company Strata Decision Technology [10]. In European hospitals, 7.98% to 16.67% of patients experienced a prolonged length of hospital stay [11–13]. In Asia, studies have shown that 5.4% to 30.5% of patients had prolonged hospital stays [14–16], and in Africa, 24.7% to 63% of patients experienced prolonged hospital stays [17–19].

Hence, hospitals face significant challenges related to insufficient surgical bed capacity and inconsistent occupancy rates in surgical units, resulting in frequent last-minute cancellations of surgeries [20]. This has a great impact on patients, their families, and health services [1,6,13]. However, it should be considered that patients who stay longer than necessary or shorter than is actually needed have an impact on the price and standard of care delivered [21]. Although patients who stayed for a prolonged time had poor care outcomes [22,23], they faced financial burdens [24,25], made it difficult for the hospital to run at a high level of efficiency [26,27], were at higher risk of developing complications [28,29], and had increased mortality [11]. Generally, prolonged hospital stays are not beneficial for either patients or hospitals [6,30].

Studies have revealed that factors such as older age, preoperative hypoalbuminemia, ASA class 3 or 4, duration of operation, in-hospital complications, comorbidity, and reoperation were factors independently associated with prolonged hospital stays [19,22,25,31,32]. However, there is a lack of consistency in what influences patients’ length of stay, particularly in surgical wards [33]. On the other hand, this research topic covers a number of previously unexplored aspects that have attracted research interest, such as postponed surgery, preoperative anemia, functional status, and source of referral [18,19], which should be investigated further. An investigation of these factors is necessary to reduce unnecessary stays in the hospital.

In an effort to enhance patient care quality and lower hospital costs, many parts of the health system are working hard to reduce patients’ unnecessary hospital stays [21,34] through interdisciplinary or multidisciplinary care, medication management, and discharge planning tools [33,34]. As in other countries, the Federal Ministry of Health (FMOH) is trying to improve the quality of surgical care by launching a strategy to save lives through safe surgery and has implemented the World Health Organization’s surgical safety checklist, which has an effect on reducing the length of hospital stay [35,36].

Although the length of stay (LOS) among patients admitted to surgical units has been declining in Western nations, it remains high in Africa [17–19,37]. This disparity is a result of the varied healthcare systems and clinical practices [38], and it may also be due to the lack of utilization of strategies implemented in hospitals related to surgical care, which influences the length of hospital stays. Moreover, there are inconsistent findings about the efficacy of LOS-reduction measures such as discharge planning tools, which are usually utilized by healthcare systems [34]. In addition, there is little published literature on factors that influence the length of hospital stays among surgical inpatients in Africa and Ethiopia. Therefore, this study aimed to assess the length of hospital stay and associated factors among adult surgical patients admitted to a surgical ward in Amhara Regional State comprehensive specialized hospitals, Ethiopia. The results of these findings provide an additional clue for the Ethiopian healthcare system to gain a better understanding of factors contributing to prolonged hospital stays with surgical care services.

## Materials and methods

### Study design, period and setting

An institutional-based cross-sectional study was conducted in the adult surgical ward in Amhara Regional State Comprehensive Specialized Hospitals from April 17 to May 22, 2023. The University of Gondar, Dessie, Felege Hiwot, and Tibebe Ghion are the four selected comprehensive specialized hospitals found in the Amhara Regional States. The University of Gondar is one of the comprehensive specialized hospitals found in the Amhara Regional State. This hospital serves more than 5 million people in its catchment area. There are more than 906 (463 nurses) healthcare professionals and 40 beds in the surgical ward [39]. The average monthly surgical patient admission was 106.

Tibebe Ghion and Felege Hiwot comprehensive specialized hospitals are found in Bahir Dar, the capital of Amhara Regional State, which is 565 kilometers away from Addis Ababa and serves more than 5 million people in each hospital. They have 105 and 66 beds in the surgical ward, respectively [40]. The average monthly surgical patient admissions were 335 and 168, respectively.

Dessie Comprehensive Specialized Hospital, found in northeastern Ethiopia. The Dessie city administration is found far from Bahir Dar (481 kilometers), which is the capital city of Amhara regional state, and 401 kilometers from Addis Ababa, the capital city of Ethiopia. The Dessie Comprehensive hospital serves a population of 5 million, has 603 health care workers, and has 66 beds in the surgical ward [41]. The average monthly surgical patient admission was 141.

### Population

All adult (≥18 years) surgical patients who underwent surgery and were admitted to a surgical ward in Amhara Regional State Comprehensive Specialized Hospitals were the source population, and all adult (≥18 years) surgical patients who underwent surgery and were admitted to a surgical ward in selected Amhara Regional State Comprehensive Specialized Hospitals during the study period were the study population.

### Inclusion and exclusion criteria

All sampled adult surgical patients who underwent both elective and emergency surgery and were admitted to a surgical ward in selected Amhara Regional State CSHs were included. The study excluded patients who were unable to communicate because of their illness and orthopedic patients.

### Sample size determination

The sample size was determined using a single population proportion formula with the following assumptions:

n = sample size

Z α/2= significance level =95% CI=1.96, d = margin of error of 0.05, and proportion of prolonged length of hospital stay from a previous study at Jimma Hospital (P), 25.3% [18].

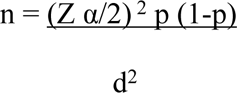

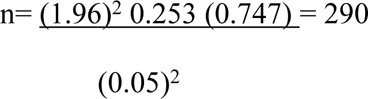

By considering 1.5 design effects and adding a 5% (22) nonresponse rate, the total sample size was 457. The sample sizes for factors were also calculated by using EpiInfo version 7.2. Based on the results, the proportion of patients with a prolonged length of hospital stay was used for the final sample size.

### Sampling procedure

From eight comprehensive specialized hospitals in the Amhara Regional States, four were selected by lottery, such as the University of Gondar, Dessie, Felege Hiwot, and Tibebe Ghion comprehensive specialized hospitals. The total number of patients who underwent surgery and were admitted to a surgical ward in those hospitals in the last month before data collection was 750. Then, based on their population sizes, the sample size was proportionally distributed among them using the proportional allocation formula. After that, eligible study participants were enrolled using a systematic random sampling technique. Then, the lottery method was employed to identify the first study participant to interview, and then every other study participant was selected until the needed sample size was obtained.

### Study variables

#### Dependent variable

Length of hospital stays (Prolonged, Normal)

#### Independent variables

Sociodemographic factors (age, sex, marital status, educational status, residency, occupation, economic status, and health insurance)

Admission-related factors (type of surgery admission (elective or emergency), time of admission, and source of referral)

Clinical factors (comorbidity, type of surgical procedure, in-hospital complications, duration of surgery (in minutes), reoperation, postponed operations, preoperative anemia, body mass index and functional status)

Behavioral factors (smoking status and alcohol status)

### Operational definitions

**Length of hospital stay:** described as the length of a single hospitalization, which is determined by subtracting the day of discharge from the day of admission.

**Prolonged LOS:** defined as an LOS greater than or equal to the 75^th^ percentile in hospitals for the entire study population that was at least 14 days in this study (Prolonged coded as 1, Normal coded as 0) [11].

**Normal LOS:** defined as LOS less than the 75^th^ percentile for the entire study population

**In-hospital complications:** The occurrence of new medical issues upon admission that were seen as negative incidents arising from the course of care and therapy rather than the course of the disease, such as pressure injuries, falls, healthcare-associated infections (HAIs), surgical complications, respiratory complications, cardiac complications, venous thromboembolism, renal failure, gastrointestinal bleeding, and adverse drug events. HAIs were further classified into specific types, such as central-line associated bloodstream, catheter-associated urinary tract infection, hospital-acquired pneumonia, ventilator-associated pneumonia, and surgical site infections [42].

**Preoperative anemia:** defined as hemoglobin (Hb) levels below 12.0 g/dL in women and 13.0 g/dL in males, was considered anemia, according to the World Health Organization (WHO) [43].

**The duration of surgery:** was defined as the time from the first incision to wound closure [44].

### Data collection tools and procedures

The data were collected from adult surgical patients admitted to a surgical ward using a pretested, structured interviewer-administered questionnaire, patient chart review, and direct measurement. The data collection instrument was adapted from previously published studies [17–19], whereas wealth index assessment questions were adopted from EDHS 2016 [45], and the functional status assessment tool was adopted from the Katz Index of Assessing Tool [46]. The questionnaire had sociodemographic information, wealth index assessment questions, admissions-related characteristics, clinical variables, behavioral information, and the Katz Index of functional status assessment tools. The data collectors were four BSc nurse professionals and three MSc nurse supervisors. The collected data were checked daily for consistency and accuracy.

### Data quality control measures

The questionnaires were translated into Amharic and back into English to maintain consistency. In this study, two senior surgical clinicians and two researchers checked for face validity. One-day training was given for data collectors and supervisors by the principal investigator on how to collect data, the objective of the study, contents of the questionnaire, timely collection, ethical issues, and interviewing technique. Continuous supervision was provided on the spot by the principal investigator and supervisors. The questionnaire was pretested on 5% (23) of the sample at Debre Tabor Comprehensive Specialized Hospital (DTCSH) before actual data collection to check acceptability and consistency. Depending on the result of the pretest, necessary modifications were made to ensure the clarity and completeness of the questionnaire. Finally, before beginning entry into EpiData version 4.6.0, the data were cleared, categorized, compiled, and checked for completeness and accuracy.

### Data processing and analysis

EpiData version 4.6.0 was used to code and enter data, which were exported to STATA version 14 (StataCorp, College Station, TX, USA) for analysis. Descriptive statistics such as frequency and percentage were used to summarize the study population. Continuous variables were expressed using measures of central tendency and variability, such as the mean, median and interquartile range (IQR). A binary logistic regression model was fitted. A variable with a p value < 0.25 was used to select potential candidates for multivariable analysis, and variables with a p value < 0.05 were considered significantly associated with the outcome variable. An odds ratio with a 95% CI was used as a measure of association. The results are presented in tables and graphs. The presence of multicollinearity among independent variables was checked by the variance inflation factor (VIF = mean 1.54). The Hosmer‒Lemeshow tests indicated that the model fit the data well (p = 0.75).

### Ethical consideration

Ethical clearance was obtained from the ethical review committee of the nursing school on behalf of the Institutional Review Board (IRB) of the University of Gondar with Ref. Number (S/N/175/2015). A support letter of cooperation was written to the Amhara Public Health Institute and comprehensive specialized hospitals of Amhara Regional States. Then, permission was obtained to undertake the study from the administrative offices of comprehensive specialized hospitals in the Amhara Regional States. All the study participants received written informed consent. The data collectors provided a brief overview of the study’s primary aim. To ensure anonymity, respondent names and other identifiable information were not included. Finally, it was made clear to study participants that their answers to private questions were confidential, and they were free to end the interview at any time and not respond to any questions. Password protection and proper disposal were used to protect the acquired data.

## Results

### Sociodemographic characteristics of the participants

In this study, 452 adult surgical patients participated, with a response rate of 99%. Approximately two-thirds, 63.05% (285), were male, and the median age was 38 years old, with nearly one-third, 29.42% (133), being within the age range of 25–34 years old. Most, 76.8% (347), were married, and more than half, 57.1% (258), were rural dwellers. Around half 47.35% (214) of the participants were farmers, and one-third of the study participants 33.85% (153) were within the lower wealth index. Three-fourths, 75% (339), had health insurance (Table 1).

**Table 1.**
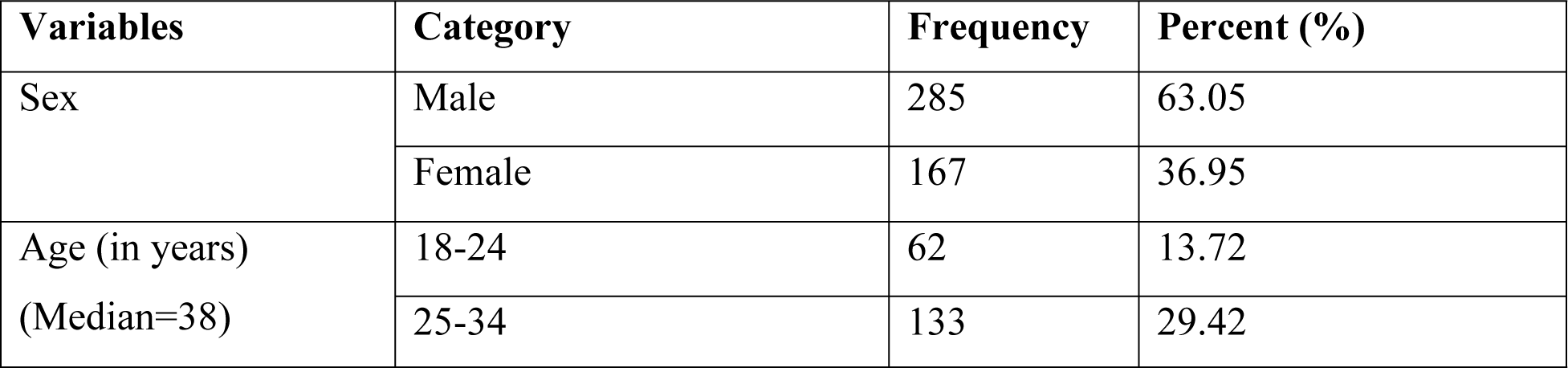
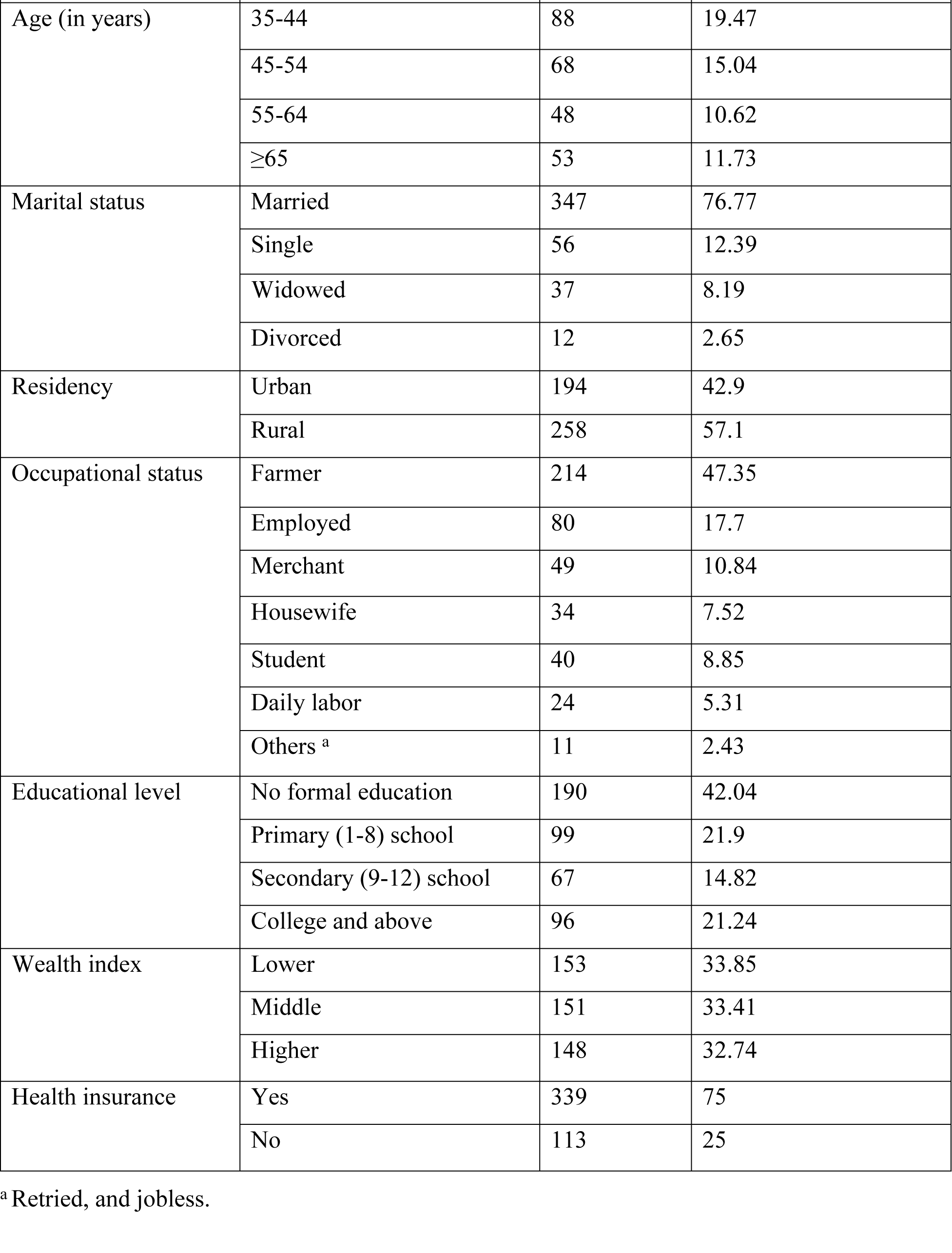
Sociodemographic characteristics of adult surgical patients admitted to a surgical ward in Amhara Regional State Comprehensive Specialized Hospitals, Ethiopia, 2023 (n = 452).

### Admission-related characteristics

Among 452 study participants, nearly two-thirds 63.72% (288) had undergone elective surgery. The majority, 82.74% (374), admitted during the day, and 72.35% (327) presented directly from their home (Table 2).

**Table 2.** Admission-related characteristics of adult surgical patients admitted to a surgical ward in Amhara Regional State Comprehensive Specialized Hospitals, Ethiopia, 2023 (n=452).

| Variables | Category | Frequency | Percent (%) |
| --- | --- | --- | --- |
| Type of surgery admission | Emergency | 164 | 36.28 |
|  | Elective | 288 | 63.72 |
| Time of admission | Day time | 374 | 82.74 |
|  | Night time | 78 | 17.26 |
| Source of referral | Home | 327 | 72.35 |
|  | Public health institution | 105 | 23.23 |
|  | Private health institution | 20 | 4.42 |

### Clinical characteristics

Out of 452 study participants, 4.65% (21) had postponed surgery, and 11.06% (50) had comorbid conditions. Two-thirds, 68.14% (308), underwent general surgery, followed by urologic procedures, 19.69% (89). During their hospital stay, 7.96% (36) developed hospital-acquired pneumonia (HAP), and 3.76% (17) experienced surgical site infection (SSI). More than one-third (38.94%, 176) of the participants had surgeries that lasted ≥110 minutes, and 22.57% (102) had preoperative anemia. Furthermore, 17.92% (81) had a low body mass index, and more than half, 51.33% (232), of them were partially dependent on their functional status (Table 3).

**Table 3.**
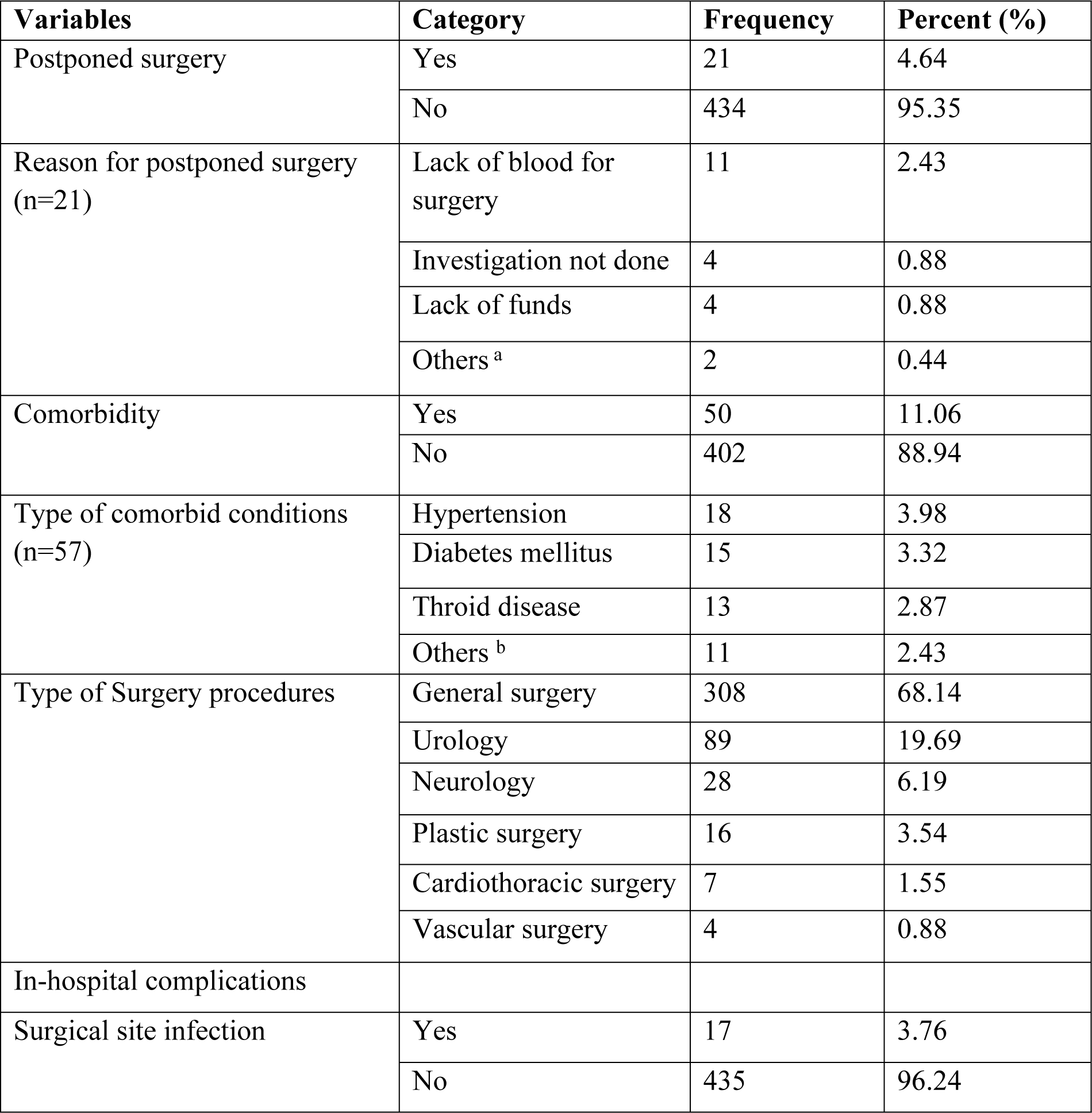
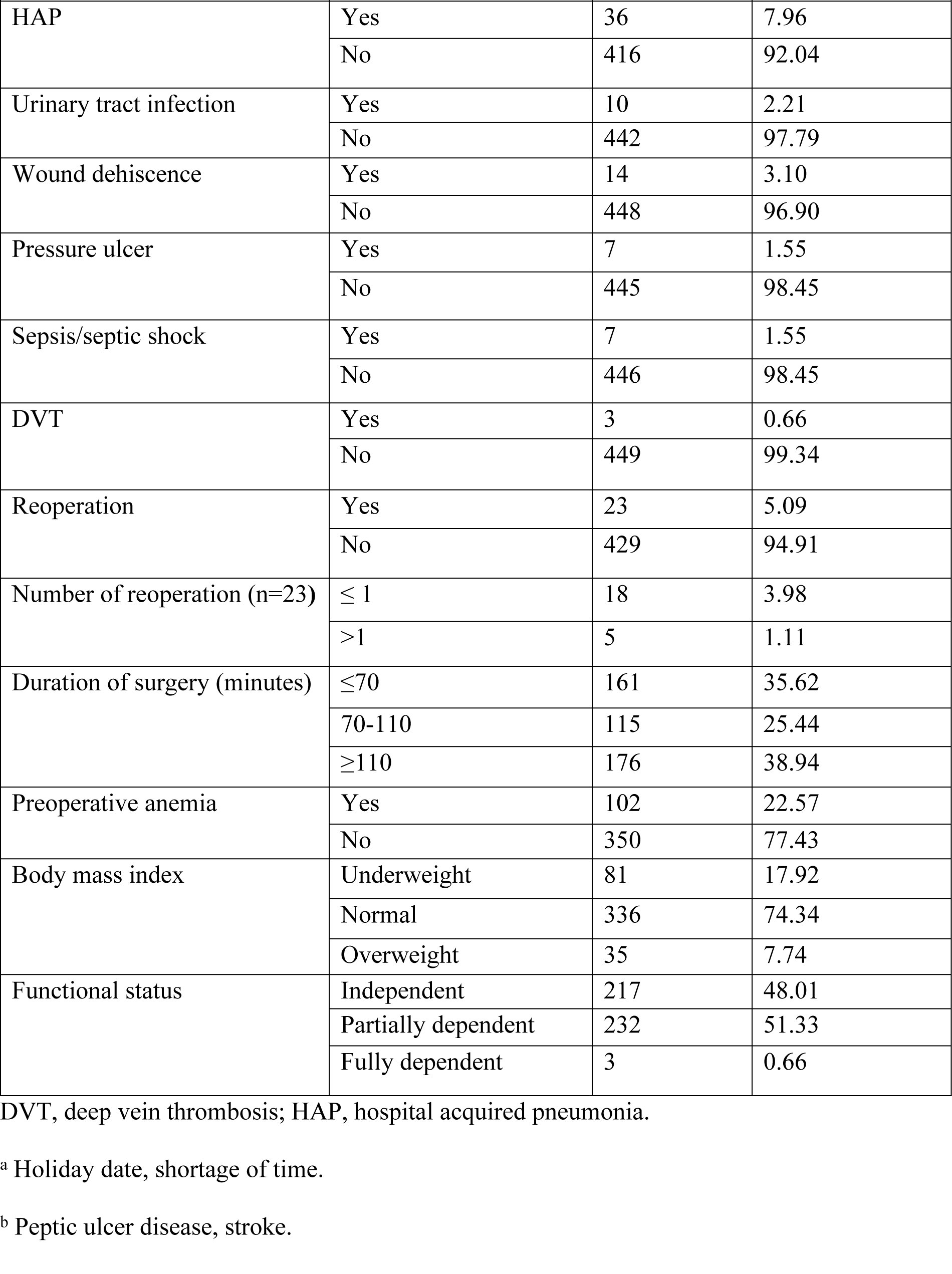
Clinical features of adult admitted surgical patients in Amhara Regional State Comprehensive Specialized Hospitals, Ethiopia, 2023 (n = 452).

| **Variables** | **Category** | **Frequency** | **Percent (%)** |
| --- | --- | --- | --- |
| Postponed surgery | Yes | 21 | 4.64 |
|  | No | 434 | 95.35 |
| Reason for postponed surgery (n=21) | Lack of blood for surgery | 11 | 2.43 |
|  | Investigation not done | 4 | 0.88 |
|  | Lack of funds | 4 | 0.88 |
|  | Others <sup>a</sup> | 2 | 0.44 |
| Comorbidity | Yes | 50 | 11.06 |
|  | No | 402 | 88.94 |
| Type of comorbid conditions (n=57) | Hypertension | 18 | 3.98 |
|  | Diabetes mellitus | 15 | 3.32 |
|  | Throid disease | 13 | 2.87 |
|  | Others <sup>b</sup> | 11 | 2.43 |
| Type of Surgery procedures | General surgery | 308 | 68.14 |
|  | Urology | 89 | 19.69 |
|  | Neurology | 28 | 6.19 |
|  | Plastic surgery | 16 | 3.54 |
|  | Cardiothoracic surgery | 7 | 1.55 |
|  | Vascular surgery | 4 | 0.88 |
| In-hospital complications |  |  |  |
| Surgical site infection | Yes | 17 | 3.76 |
|  | No | 435 | 96.24 |
| HAP | Yes | 36 | 7.96 |
|  | No | 416 | 92.04 |
| Urinary tract infection | Yes | 10 | 2.21 |
|  | No | 442 | 97.79 |
| Wound dehiscence | Yes | 14 | 3.10 |
|  | No | 448 | 96.90 |
| Pressure ulcer | Yes | 7 | 1.55 |
|  | No | 445 | 98.45 |
| Sepsis/septic shock | Yes | 7 | 1.55 |
|  | No | 446 | 98.45 |
| DVT | Yes | 3 | 0.66 |
|  | No | 449 | 99.34 |
| Reoperation | Yes | 23 | 5.09 |
|  | No | 429 | 94.91 |
| Number of reoperation (n=23) | ≤ 1 | 18 | 3.98 |
|  | >1 | 5 | 1.11 |
| Duration of surgery (minutes) | ≤70 | 161 | 35.62 |
|  | 70-110 | 115 | 25.44 |
|  | ≥110 | 176 | 38.94 |
| Preoperative anemia | Yes | 102 | 22.57 |
|  | No | 350 | 77.43 |
| Body mass index | Underweight | 81 | 17.92 |
|  | Normal | 336 | 74.34 |
|  | Overweight | 35 | 7.74 |
| Functional status | Independent | 217 | 48.01 |
|  | Partially dependent | 232 | 51.33 |
|  | Fully dependent | 3 | 0.66 |

DVT, deep vein thrombosis; HAP, hospital acquired pneumonia.
<sup>a</sup> Holiday date, shortage of time.
<sup>b</sup> Peptic ulcer disease, stroke.

### Behavioral characteristics of the study participants

Among the study participants, 41.59% (188) currently consumed alcohol, whereas the majority, 90.71% (410) of the respondents, reported that they were never cigarette smokers (Table 4).

**Table 4.** Behavioral characteristics of adult surgical patients admitted to a surgical ward in Amhara Regional State Comprehensive Specialized Hospitals, Ethiopia, 2023 (n = 452).

| Variables | Category | Frequency | Percent (%) |
| --- | --- | --- | --- |
| Alcohol drinking status | Never | 133 | 29.2 |
|  | Current | 188 | 41.59 |
|  | Past drinker | 131 | 28.98 |
| Smoking status | Never | 410 | 90.71 |
|  | Current | 29 | 6.42 |
|  | Former | 13 | 2.88 |

### Length of hospital stay

In the current study, the prevalence of prolonged hospital stays was 26.5% (95% CI: 22.7–30.8). The average length of hospital stay was 9.38 (SD: ±7.31) days. The minimum and maximum lengths of hospital stays were 2 and 60 days, respectively (Fig 1). The highest average length of hospital stay occurred among those who underwent cardiothoracic surgery, followed by plastic surgery procedures (Fig 2).

**Fig 1.**
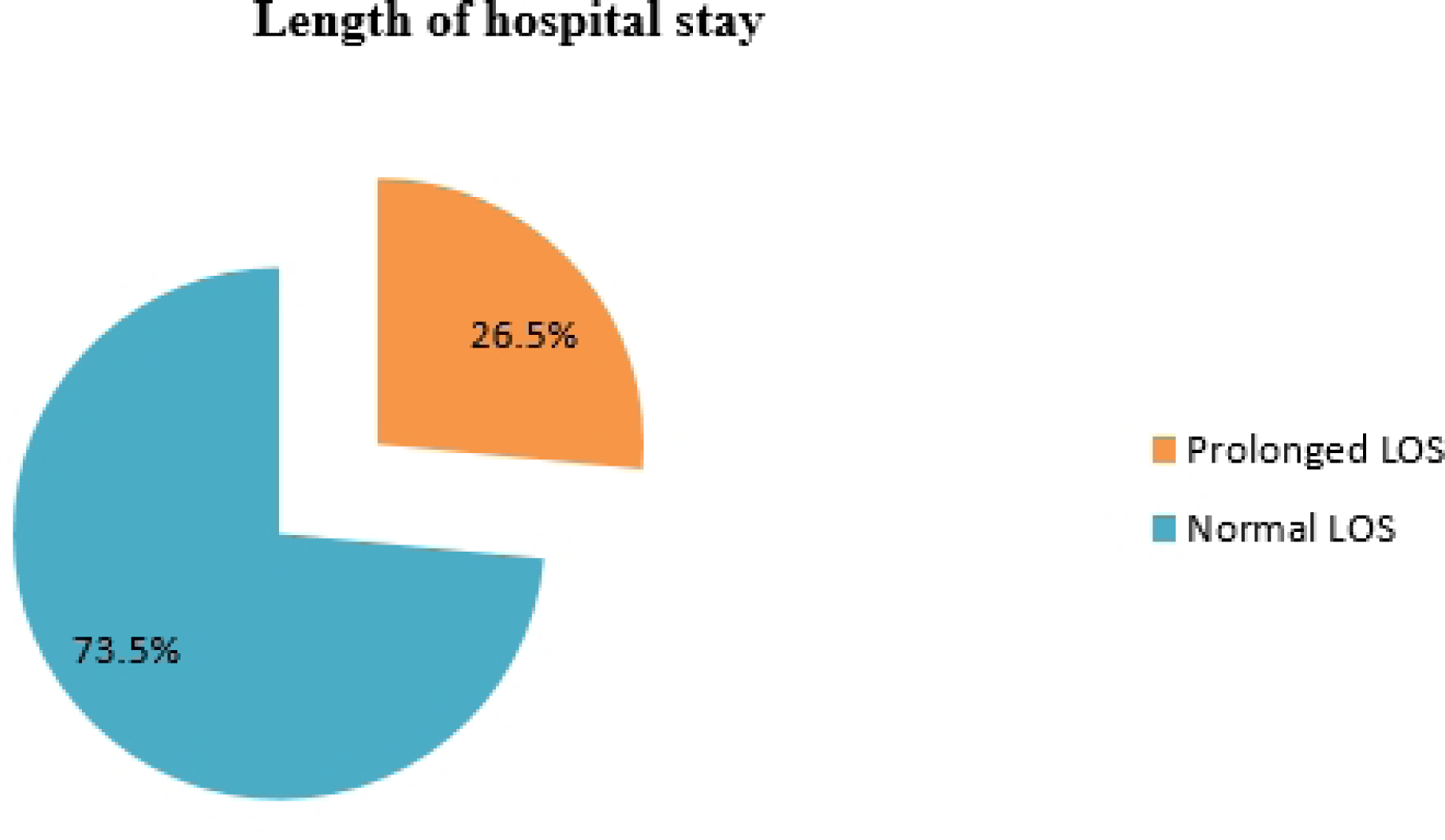
Proportion of length of hospital stays among adult surgical patients admitted to a surgical ward in Amhara Regional State Comprehensive Specialized Hospitals, Ethiopia, 2023 (n=452).

**Fig 2.**
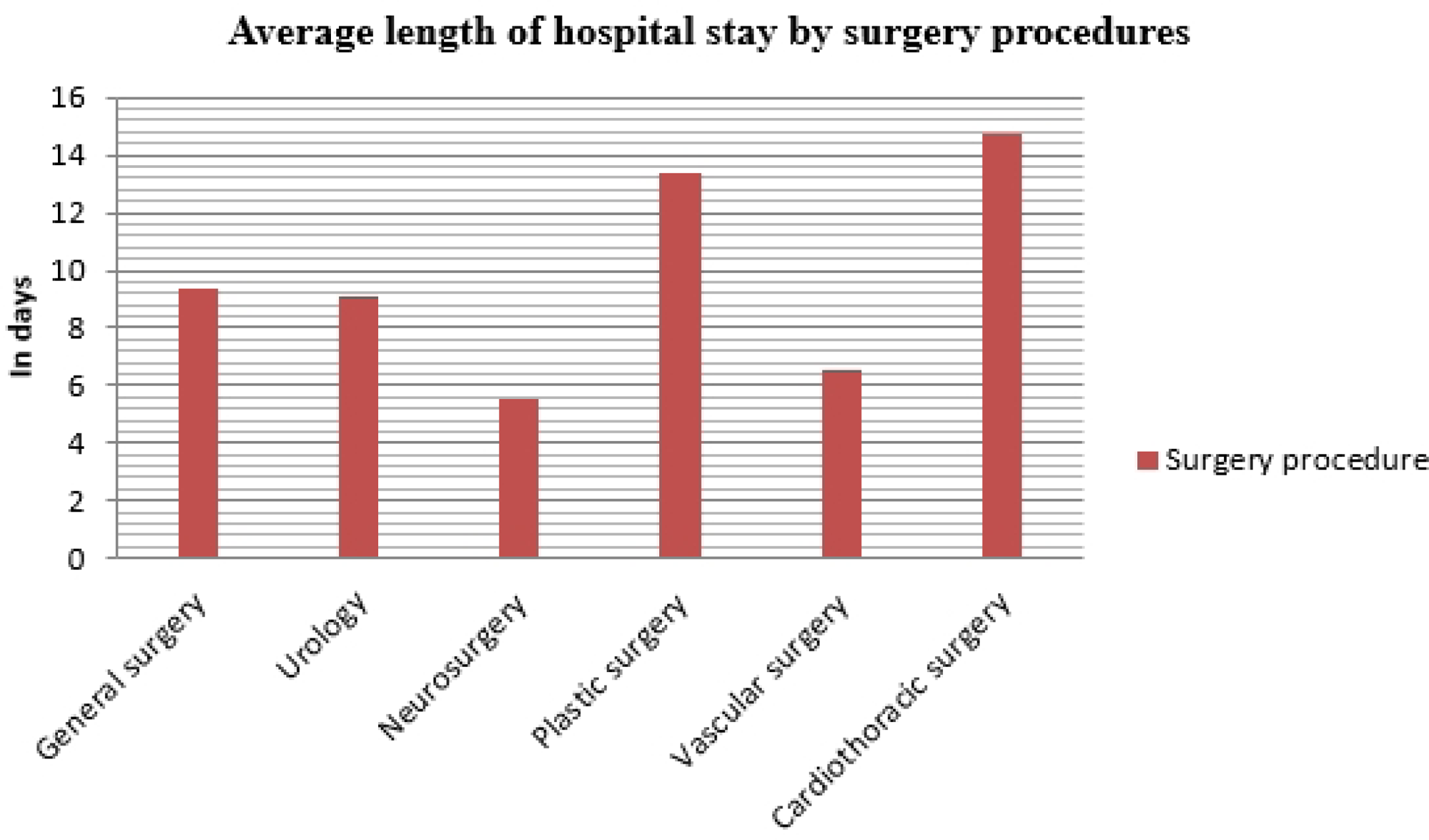
Average length of hospital stays among adult surgical patients admitted to a surgical ward in Amhara Regional State Comprehensive Specialized Hospitals, Ethiopia, 2023 (n=452).

### Factor associated with prolonged length of hospital stay

In the bivariable analysis, sex, marital status, health insurance, educational status, type of surgery admission, source of referral, time of admission, body mass index, wealth index, HAP, surgical site infection (SSI), reoperation, preoperative anemia, alcohol status, and duration of surgery were factors associated with prolonged LOS. However, only four variables, source of referral, HAP, preoperative anemia, and duration of surgery, were significantly associated with prolonged LOS in multivariable logistic regression analysis.

The odds of having prolonged LOS were 2.5 times (AOR = 2.46; 95% CI: 1.09–5.57) higher among patients referred from another public health institution compared with those presented directly from home. The odds of having prolonged LOS were 3.2 times (AOR = 3.18; 95% CI: 1.28–7.89) greater among patients who developed hospital-acquired pneumonia than among those who had no HAP. Patients who had surgery lasting ≥110 minutes were 2.5 times (AOR = 2.48; 95% CI: 1.25–4.91) more likely to have prolonged LOS than those who had an operation time of ≤70 minutes. Furthermore, the odds of having prolonged LOS were 3.4 times (AOR = 3.37; 95% CI: 1.88–6.04) higher among preoperative anemia patients than among those who had no preoperative anemia (Table 5).

**Table 5.**
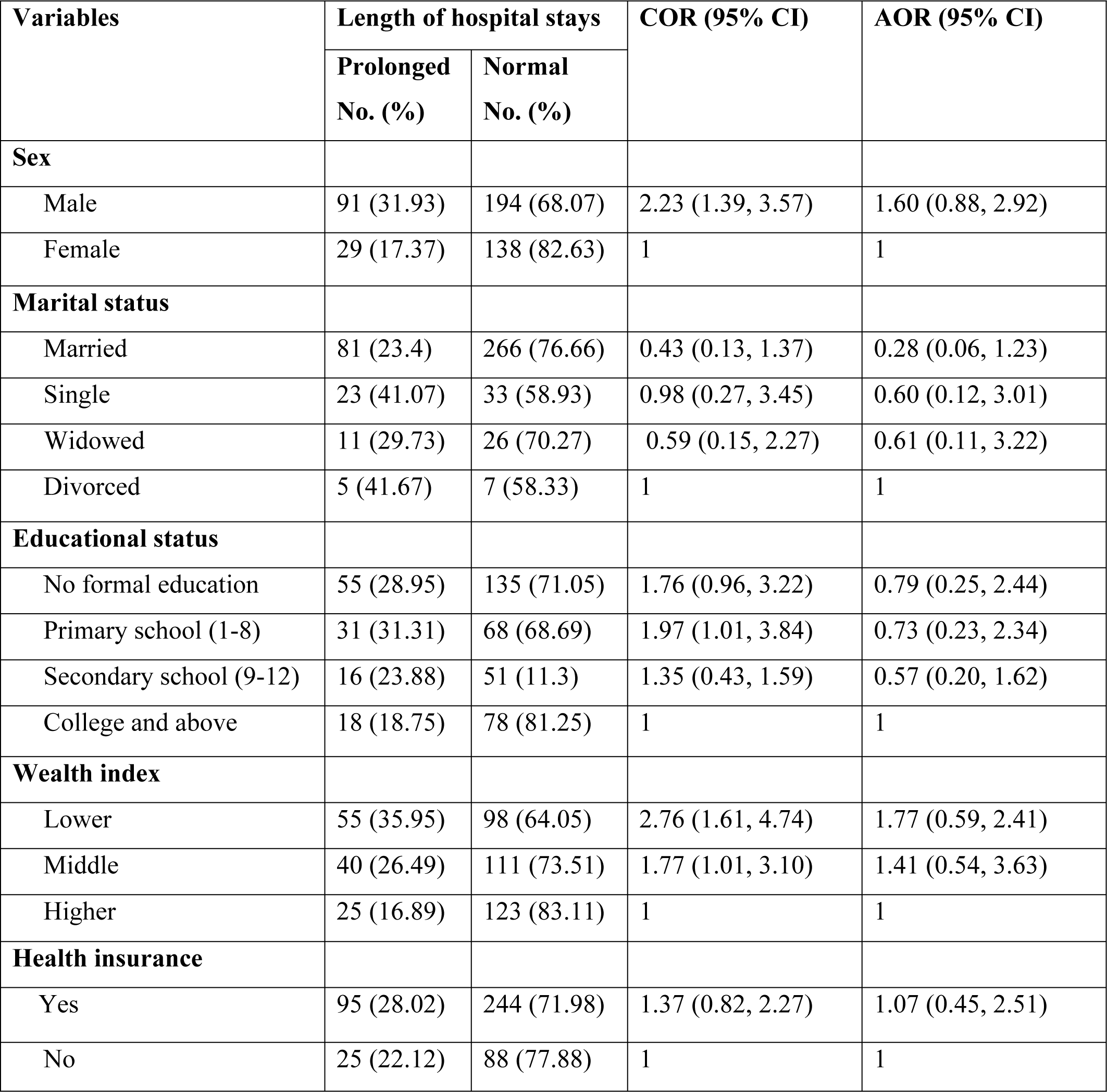
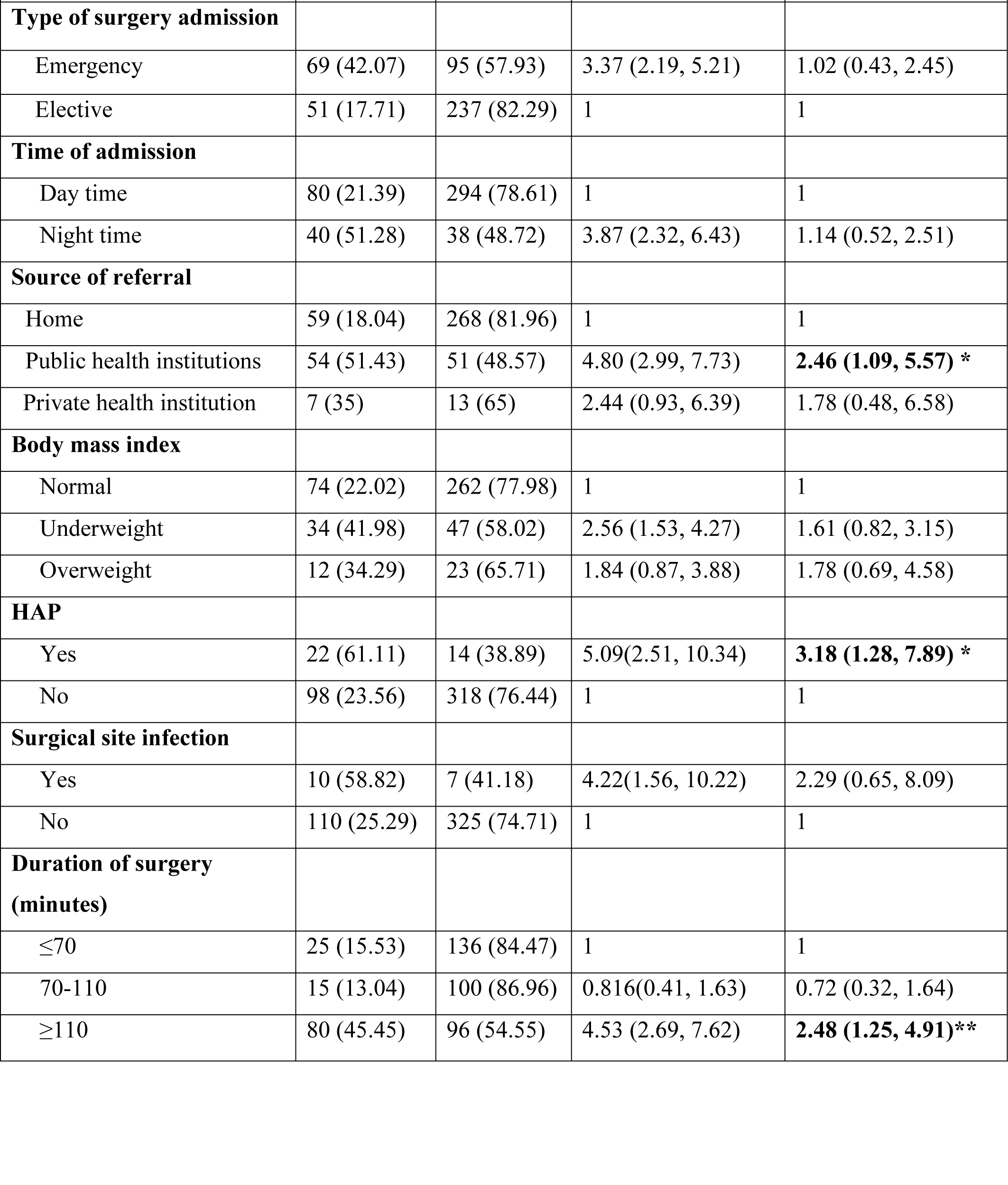
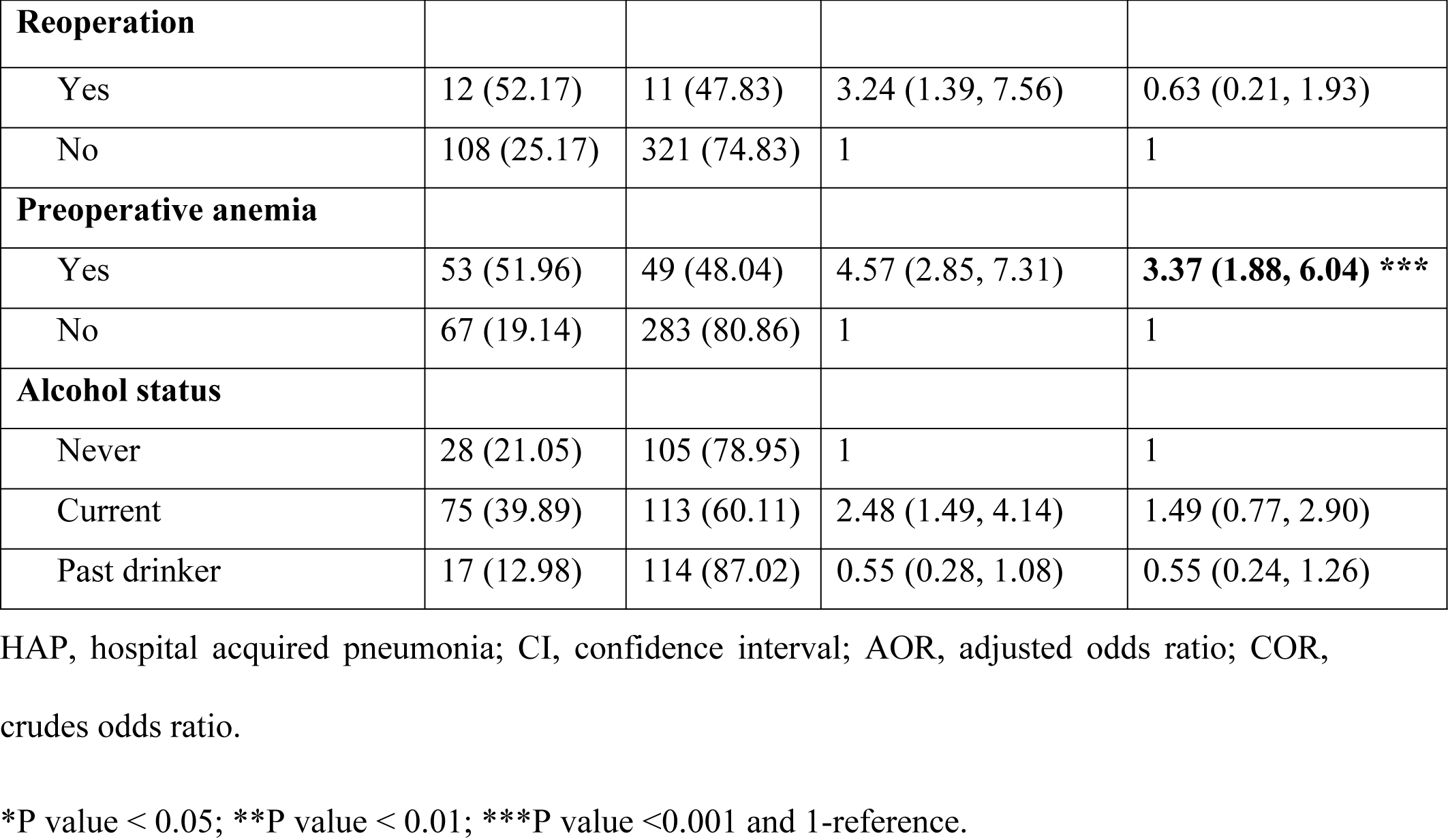
Bivariable and multivariable analysis of factors associated with the length of hospital stay among adult surgical patients admitted to a surgical ward in Amhara Regional State Comprehensive Specialized Hospital, Ethiopia, 2023 (n = 452).

| Variables | Length of hospital stays |  | COR (95% CI) | AOR (95% CI) |
| --- | --- | --- | --- | --- |
|  | Prolonged<br>No. (%) | Normal<br>No. (%) |  |  |
| **Sex** |  |  |  |  |
| Male | 91 (31.93) | 194 (68.07) | 2.23 (1.39, 3.57) | 1.60 (0.88, 2.92) |
| Female | 29 (17.37) | 138 (82.63) | 1 | 1 |
| **Marital status** |  |  |  |  |
| Married | 81 (23.4) | 266 (76.66) | 0.43 (0.13, 1.37) | 0.28 (0.06, 1.23) |
| Single | 23 (41.07) | 33 (58.93) | 0.98 (0.27, 3.45) | 0.60 (0.12, 3.01) |
| Widowed | 11 (29.73) | 26 (70.27) | 0.59 (0.15, 2.27) | 0.61 (0.11, 3.22) |
| Divorced | 5 (41.67) | 7 (58.33) | 1 | 1 |
| **Educational status** |  |  |  |  |
| No formal education | 55 (28.95) | 135 (71.05) | 1.76 (0.96, 3.22) | 0.79 (0.25, 2.44) |
| Primary school (1-8) | 31 (31.31) | 68 (68.69) | 1.97 (1.01, 3.84) | 0.73 (0.23, 2.34) |
| Secondary school (9-12) | 16 (23.88) | 51 (11.3) | 1.35 (0.43, 1.59) | 0.57 (0.20, 1.62) |
| College and above | 18 (18.75) | 78 (81.25) | 1 | 1 |
| **Wealth index** |  |  |  |  |
| Lower | 55 (35.95) | 98 (64.05) | 2.76 (1.61, 4.74) | 1.77 (0.59, 2.41) |
| Middle | 40 (26.49) | 111 (73.51) | 1.77 (1.01, 3.10) | 1.41 (0.54, 3.63) |
| Higher | 25 (16.89) | 123 (83.11) | 1 | 1 |
| **Health insurance** |  |  |  |  |
| Yes | 95 (28.02) | 244 (71.98) | 1.37 (0.82, 2.27) | 1.07 (0.45, 2.51) |
| No | 25 (22.12) | 88 (77.88) | 1 | 1 |
|  | Prolonged<br>No. (%) | Normal<br>No. (%) |  |  |
| **Type of surgery admission** |  |  |  |  |
| Emergency | 69 (42.07) | 95 (57.93) | 3.37 (2.19, 5.21) | 1.02 (0.43, 2.45) |
| Elective | 51 (17.71) | 237 (82.29) | 1 | 1 |
| **Time of admission** |  |  |  |  |
| Day time | 80 (21.39) | 294 (78.61) | 1 | 1 |
| Night time | 40 (51.28) | 38 (48.72) | 3.87 (2.32, 6.43) | 1.14 (0.52, 2.51) |
| **Source of referral** |  |  |  |  |
| Home | 59 (18.04) | 268 (81.96) | 1 | 1 |
| Public health institutions | 54 (51.43) | 51 (48.57) | 4.80 (2.99, 7.73) | **2.46 (1.09, 5.57) \*** |
| Private health institution | 7 (35) | 13 (65) | 2.44 (0.93, 6.39) | 1.78 (0.48, 6.58) |
| **Body mass index** |  |  |  |  |
| Normal | 74 (22.02) | 262 (77.98) | 1 | 1 |
| Underweight | 34 (41.98) | 47 (58.02) | 2.56 (1.53, 4.27) | 1.61 (0.82, 3.15) |
| Overweight | 12 (34.29) | 23 (65.71) | 1.84 (0.87, 3.88) | 1.78 (0.69, 4.58) |
| **HAP** |  |  |  |  |
| Yes | 22 (61.11) | 14 (38.89) | 5.09(2.51, 10.34) | **3.18 (1.28, 7.89) \*** |
| No | 98 (23.56) | 318 (76.44) | 1 | 1 |
| **Surgical site infection** |  |  |  |  |
| Yes | 10 (58.82) | 7 (41.18) | 4.22(1.56, 10.22) | 2.29 (0.65, 8.09) |
| No | 110 (25.29) | 325 (74.71) | 1 | 1 |
| **Duration of surgery**<br>**(minutes)** |  |  |  |  |
| ≤70 | 25 (15.53) | 136 (84.47) | 1 | 1 |
| 70-110 | 15 (13.04) | 100 (86.96) | 0.816(0.41, 1.63) | 0.72 (0.32, 1.64) |
| ≥110 | 80 (45.45) | 96 (54.55) | 4.53 (2.69, 7.62) | **2.48 (1.25, 4.91)\*\*** |

**Table 5.** (Continued)
| Variables | Length of hospital stays |  | COR (95% CI) | AOR (95% CI) |
| --- | --- | --- | --- | --- |
|  | Prolonged<br>No. (%) | Normal<br>No. (%) |  |  |
| **Reoperation** |  |  |  |  |
| Yes | 12 (52.17) | 11 (47.83) | 3.24 (1.39, 7.56) | 0.63 (0.21, 1.93) |
| No | 108 (25.17) | 321 (74.83) | 1 | 1 |
| **Preoperative anemia** |  |  |  |  |
| Yes | 53 (51.96) | 49 (48.04) | 4.57 (2.85, 7.31) | **3.37 (1.88, 6.04) \*\*\*** |
| No | 67 (19.14) | 283 (80.86) | 1 | 1 |
| **Alcohol status** |  |  |  |  |
| Never | 28 (21.05) | 105 (78.95) | 1 | 1 |
| Current | 75 (39.89) | 113 (60.11) | 2.48 (1.49, 4.14) | 1.49 (0.77, 2.90) |
| Past drinker | 17 (12.98) | 114 (87.02) | 0.55 (0.28, 1.08) | 0.55 (0.24, 1.26) |

HAP, hospital acquired pneumonia; CI, confidence interval; AOR, adjusted odds ratio; COR, crudes odds ratio.
\*P value < 0.05; \*\*P value < 0.01; \*\*\*P value <0.001 and 1-reference.

## Discussion

Assessing the length of hospital stay among surgical patients admitted to the surgical ward is one of the most helpful approaches to assess the quality of surgical treatment given to patients. Additionally, identifying patients who have prolonged hospital stays could be a useful strategy for lowering the unnecessary length of stay (LOS) in hospitals.

According to the findings of the current study, 26.5% experienced prolonged LOS. This finding is consistent with previous studies done in Oman (30.5) [16], Boston (27.7%) [47], and Jimma University Medical Center (JUMC) (25.3%) [18]. However, higher than the study conducted in Australia (9.7%) [11], the Philippines (19.17%) [15], the USA (22%) [48], 18 Polish and German surgical centers (4.92%) [49], and Saudi Arabia (21%) [50]. This discrepancy may be due to advancements in the health care system and the fact that most of the care is provided by specialist healthcare professionals in developed countries, which is difficult in low- and middle-income countries. Moreover, in 18 Polish and German surgical centers, the difference may be due to patients who had at least 8 days of hospital stay, which was considered prolonged LOS [49]. Studies have shown that there are numerous barriers that developing countries face in providing high-quality surgical care [51].

On the other hand, this study result is lower than the study conducted in Nigeria (63%) [17] and Thailand (54.9%) [52]. In the case of Nigeria, this discrepancy may be due to the study period and the lower cutoff points used, which were at least 7 days, which is less than the cutoff point used in this study. In Thailand, the difference may be due to the source population; the study was performed on patients who had a fractured neck of the femur. According to a study, fractured patients had a longer hospital stay than other surgical patients [53].

The current study revealed that the source of referral, preoperative anemia, HAP, and duration of surgery had a statistically significant association with prolonged LOS. This study found that referred patients from another public health institution were more likely to have a prolonged hospital stay than those presented directly from home. This finding is supported by previous studies conducted in Australia and the USA [28,54]. There was no clear reason why referred patients experienced prolonged LOS. However, a possible explanation may be that patients who have been referred may need more time and resources to treat because they may be sicker than patients who have not been referred. As a result, these factors increased their treatment durations and investigations, which in turn increased their length of hospital stay. Additionally, the length of hospital stays is increased by inadequate systems for exchanging patient information among hospitals when referring [55]. A key area emerging from the current results is that the source of referral is one of the factors that leads to prolonged hospital stays.

The present study found that patients who had preoperative anemia were 3.4 times more likely to have a prolonged hospital stay than those without preoperative anemia. This result is in line with the studies conducted in Singapore and Germany [31,43]. The possible reason may be due to untreated preoperative anemia related to complications following a surgical procedure as well as a higher demand for blood transfusion and hence prolonged LOS [31]. An additional explanation may be that patients with anemia frequently experience fatigue and dizziness, which can make it difficult for patients to participate in their own therapy. As a result, this may extend the hospital stay. Studies have shown that early detection of anemic patients is essential in the context of surgical planning (at least 2-4 weeks prior to surgery). In emergency cases, parenteral substitution with iron as directed at the end of surgery or postoperatively, especially in cases of perioperative blood loss of more than 500 ml, can improve treatment outcomes [43]. This finding suggests that having preoperative anemia tends to extend hospital stays for patients.

This study revealed that patients who had hospital-acquired pneumonia were 3.2 times more likely to have prolonged LOS than those who had no HAP. This finding is consistent with a study conducted in the USA [47]. The possible explanation may be related to the fact that the innate immune system predominantly directs its cells to fight a lung infection and has a delayed response to wound healing, which in turn leads to an increased duration of treatment and hospital stay [56]. The result holds implications for the presence of hospital-acquired pneumonia and how it impacts the length of hospital stays for patients.

The current study found that patients who had a duration of surgery lasting ≥110 minutes were 2.5 times more likely to have a prolonged LOS than those who had a duration of surgery ≤70 minutes. This result is supported by several previous studies conducted in Japan, China, and the USA [22,32,57]. The possible reason may be that an increased operating duration is usually related to a more complicated case, a higher risk of intraoperative complications, and an increase in estimated blood loss (EBL). These factors may in turn affect the patient’s recovery and result in a prolonged hospital stay [22]. This finding suggests that having prolonged surgery is one of the significant factors that increases the duration of hospital stays. The limitations of this study were biased toward social desirability and recall bias. These were minimized by a detailed explanation of the study’s aims, used anonymously, interviews conducted separately, and chart review.

## Conclusion

This study found a significant proportion of prolonged hospital stays. Source of referral, preoperative anemia, duration of surgery, and hospital-acquired pneumonia were factors associated with a prolonged hospital stay. Therefore, strengthening the established information system among hospitals is better for exchanging patient information when referring patients. Ensuring that referred patients have received necessary treatments and investigations at hospitals. Screening and treating patients for anemia is advisable before conducting surgery. Healthcare providers should use standard safety checklists and precautions for infection prevention to enhance the quality of care. Future researchers should consider additional hospital-related factors and factors that can be controlled by the healthcare system and use mixed methods of study to determine more factors associated with prolonged hospital stays.

## Data Availability

All relevant data are within the manuscript

## Acronyms

AOR: adjusted odds ratio
BMI: body mass index
CI: confidence interval
CSHs: comprehensive specialized hospitals
DCSH: Dessie Comprehensive Specialized Hospital
DTCSH: Debre Tabor Comprehensive Specialized Hospital
FHCSH: Felege Hiwot Comprehensive Specialized Hospital
HAP: Hospital Acquired Pneumonia
LOS: length of stay
PLOS: prolonged length of stay
TGCSH: Tibebe Ghion Comprehensive Specialized Hospital
UoGCSH: University of Gondar Comprehensive Specialized Hospital
USA: United States of America
WHO: World Health Organization.

## Data Sharing Statement

Data are available by contacting the corresponding author.

## Consent for Publication

All the participants consented to publish the study in this journal.

## Acknowledgments

First, I would like to thank the University of Gondar, College of Medicine and Health Sciences, and the School of Nursing for providing me with the opportunity to pursue this master’s program. Moreover, I would like to thank the data collectors, supervisors and all the study participants for their time and willingness to take part in the study.

